# XAV-19 a Glyco-Humanized polyclonal antibody targeting SARS-CoV-2 accelerates the recovery of mild to moderate COVID-19 patients and keeps its neutralizing activity against Omicron and its subvariants

**DOI:** 10.1101/2023.10.09.23296726

**Authors:** Garyfallia Poulakou, Pierre-Joseph Royer, Nikolai Evgeniev, Gwénaëlle Evanno, Françoise Shneiker, Bernard Vanhove, Odile Duvaux, Stéphane Marot, Vincent Calvez

## Abstract

**Background:** XAV-19 is a glyco-humanized swine polyclonal antibody targeting SARS-CoV-2. The safety and clinical efficacy of XAV-19 was investigated in patients with a WHO score of 2 to 4 in the WHO 7-point ordinal scale. The activity of XAV-19 against Omicron and its subvariants was assessed *in vitro*.

**Methods:** A phase II/III, multicentric randomized double-blind placebo-controlled, clinical trial was conducted to evaluate the safety and clinical efficacy of XAV-19 in inpatients with COVID-19 requiring or not oxygen therapy and outpatients not requiring oxygen (EUROXAV trial, NCT04928430). Most patients were not vaccinated. The primary endpoint was the proportion of patients with an aggravation of COVID-19 within 8 days after treatment. Binding and neutralization of Omicron or its subvariants by XAV-19 was investigated by ELISA or with a whole virus neutralization assay.

**Results:** Patients received either 150mg of XAV-19 (N=139) or placebo (N=140). Low enrolment forced the premature trial termination. XAV-19 was well tolerated. No difference in the primary endpoint, nor in the proportion with an improvement at day 8 (secondary endpoint) was observed between the 2 groups. For patients not requiring oxygen therapy, XAV-19 reduced the time to improvement significantly (7 days *vs* 14 days p=0.0159). Neutralizing activity against Omicron and BA.2, BA2.12.1, BA.4/5 and BQ1.1 subvariants was shown in vitro.

**Conclusions:** XAV-19 did not reduce the aggravation in COVID-19 patients. While it did not bring any benefit to patients requiring oxygen, it reduced the time to improvement for patients not requiring oxygen (WHO score 2 or 3). These preliminary clinical data might indicate a therapeutic potential for patients with mild to moderate COVID-19 requiring supplementation with anti-SARS-CoV-2 neutralizing antibodies.

## INTRODUCTION

Since the beginning of the Coronavirus disease 2019 (COVID-19) pandemic, caused by Severe Acute Respiratory Syndrome Coronavirus-2 (SARS-CoV-2), mass vaccination campaigns from December 2020, dramatically reduced the impact of SARS-CoV2, limiting the pandemic and reducing the number of deaths. Yet, prophylactic or therapeutic drugs are still needed for immunocompromised patients or people who do not respond to the vaccination (1,2).

Numerous neutralizing monoclonal antibodies (mAb) have been developed against SARS-CoV-2. Most of them were raised against the original Wuhan-type virus and see their neutralization potential abrogated or reduced against variants. Moreover, mAb may favor the emergence of SARS-CoV-2 resistant variant in immunocompromised patients (3). Also, approved anti-SARS-CoV-2 mAb are no more recommended since end 2022 since they are unlikely to be effective against emerging strains of SARS-CoV-2 (4–6). Owing to their potential to bind multiple target epitopes and maintain their neutralizing activity despite mutations, polyclonal antibodies (pAbs) from animal origin represent a promising approach against COVID-19 (7,8). Hyperimmunization of qualified and selected animals guarantees large volumes of high-titer and controlled pAb. Moreover, the whole process of production and purification is quick, affordable and compatible with clinical manufacturing. Thereby, pAbs against SARS-CoV-2 are being tested in clinical trials (9–11).

We developed XAV-19, a swine glyco-humanized polyclonal antibody (GH-pAb) directed against the Wuhan-type SARS-CoV-2 RBD (12). XAV-19 is produced in alpha 1,3-galactosyltransferase (GGTA1)/cytidine monophosphate-N-acetylneuraminic acid hydroxylase (CMAH) double KO pigs, preventing the formation of immune complexes thus avoiding post-infusion serum sickness and allergies (13). Clonal origin of the double KO pigs limits the risk for batch-to-batch variation. XAV-19 broadly neutralizes variants (14) and has been introduced in clinic since 2020 where it displayed no major safety issues in a phase IIa clinical trial for patients under oxygen (9). Considering the later publications on the benefits of anti-SARS-CoV-2 antibodies when administered early, we investigated the clinical impact of XAV-19 for patients (in- or out-) suffering from mild to moderate COVID19. The results of the EUROXAV study (NCT04928430, EudraCT: 2020-005979-12), an international, placebo-controlled, double-blind, randomized clinical trial designed to evaluate the efficacy and safety of XAV-19 in patients with moderate COVID-19 requiring or not oxygen therapy are presented. Besides, as SARS-CoV-2 keep evolving, the activity of XAV-19 against Omicron and its current spreading subvariants was tested.

## METHODS

### Phase II/III Trial

#### Study design

EUROXAV (NCT04928430; EudraCT: 2020-005979-12) was a multicenter, international phase II/III, double-blind, placebo-controlled, randomized clinical trial conducted in 14 hospitals from 5 countries (Bulgaria, Greece, Romania, Spain, Turkey) between March 2021 and October 2022. This trial followed the International Council for Harmonization E6 guideline for good clinical practice and the principles of the Declaration of Helsinki. An independent data safety monitoring board (DSMB) examined the data for safety on a regular basis during the trial.

#### Participants

Adult patients must have SARS-CoV-2-confirmed infection (RT-PCR, RT-qPCR or antigen test in the last 10 days) and present signs of respiratory disease with at least 2 clinical symptoms related to COVID-19 started less than 10 days prior to screening visit. Patients should have SpO2 > 90% (at ambient air) and require or not low-flow oxygen therapy (score 2, 3 or 4 on WHO clinical progression 7-point ordinal scale (15)). Exclusion criteria were a positive SARS-CoV-2 test > 10 days, multiorgan failure, immediate ICU hospitalization, critical respiratory illness, requirement of oxygenation at flow rate > 6L/min, or signs of severe systemic illness (respiratory rate ≥ 30 per min, heart rate ≥ 125 per min, PaO2/FiO2 < 300), participation of another trial, pregnancy or breastfeeding. Eligible patients were randomized in a 1:1 ratio to receive either XAV-19 (150mg) diluted in sterile NaCl 0.9% or placebo (NaCl 0.9% only) as a 1-hour intravenous perfusion. Randomization was stratified by center and by WHO score (WHO score = 2 or 3 versus WHO score = 4). Patients were monitored during the infusion and the following hour. On the investigator judgement, the patient was either hospitalized or discharged (ambulatory patients). Dexamethasone, antithrombotic prophylaxis, antibiotics and antiviral medication (except anti-SARS-CoV-2 mAb or convalescent plasma) were allowed according to the judgement of the investigator and to the national guidelines or standard of care.

#### Sample size calculation

Sample size calculation initially assumed 15% progression of COVID-19 in the placebo arm versus 8% in the XAV-19 arm and a two-sided 5% type I error rate. A sample of 650 patients would provide the trial with 80% power. Considering a drop-out rate of 25%, 870 patients were planned.

#### Primary endpoint

The primary endpoint was the proportion of patients with an aggravation of COVID-19 within 8 days. The aggravation was defined as a worsening of the score of at least 1 point on the WHO 7-point ordinal scale compared to the score at randomization.

#### Secondary endpoints

Secondary endpoints were proportion of patients with an aggravation of COVID-19 within 8 days after treatment (defined as a worsening of the score of at least 2 points on the WHO clinical progression 7-point ordinal scale compared to the score at randomization); proportion of patients with an aggravation/improvement of COVID-19 between day 8 and day 15 (defined as a worsening/improvement of the score of at least 1 point on the WHO 7-point ordinal scale); time to aggravation/improvement measured by the proportion of patients with an aggravation/improvement at each measurement days (3, 5, 8, 15) after randomization. Safety outcomes included the cumulative incidence of any Adverse Event (AE). A stratified log rank-test will be used to compare treatment effect between groups, after adjustment on the stratification variables at baseline.

#### Statistical analyses

The global alpha type I error with bilateral hypotheses for this study was fixed at 5%. Binary logistic regression model was performed for the primary efficacy analysis. Association between the randomization group and the progression of COVID-19 was adjusted on centre, WHO score, country, age, body mass index (BMI), gender and comorbidities (defined as BMI>30, diabetes DNID, diabetes DID, cardiac disorder, vascular disorder, hypercholesterolemia, renal failure, lung disease COPD and/or asthma). For efficacy analyses, the strategy “Missing=Failure” is performed for patients who withdraw before D8 or for patients with missing data. Under this strategy, patients without an available assessment at D8 will be considered as having an aggravation. Replacement of missing data is done in the ITT and TARGET populations. The comparisons between XAV-19 group and placebo group of two or more qualitative variables was made using the χ2 test, the continuity-corrected χ2 test or Fisher exact test, according to the expected values under the assumption of independence. Comparisons of quantitative variables between XAV-19 and placebo groups were made using a Student t-test (parametric test comparing means) or Mann-Whitney-Wilcoxon test (non-parametric test comparing ranks) depending on the distribution of the variable of interest. A log rank test was used to compare time to aggravation/improvement between randomization groups. Kaplan-Meier curves are presented by randomization groups and by WHO score at baseline.

#### Spike/ACE-2 neutralization assay

Recombinant SARS-CoV-2 RBD (1µg/ml in carbonate/bicarbonate buffer pH 9.0) were immobilized overnight at 4°C on Maxisorp microtiter plates. After washes and saturation with PBS-Tween 0.05%-BSA2%, successive dilutions of XAV-19 or tixagevimab/cilgavimab were added for 30 min, followed by ACE-2-mFc Tag ligand (Sino Biological; final concentration 125 ng/ml). After 1h incubation and washing, the ACE-2 mFc Tag was revealed using a peroxidase-conjugated anti-mouse secondary antibody. Binding intensity was revealed by addition of TMB. Reaction was stopped with 50 μl of 1M H2S04 and optical density was taken at 450 nm.

#### Cytopathogenic Effect (CPE)

Vero cells (CCL-81) and Vero E6 cells (CRL-1586) were obtained from ATCC. SARS-CoV-2 clinical isolates were isolated from SARS-CoV-2 RT-PCR positive patients and viral stocks were generated by one passage on Vero cells. Titration of viral stock was performed on Vero E6 by limiting dilution assay allowing calculation of tissue culture infective dose 50% (TCID50). Serial dilutions of XAV-19 (50μl) were incubated with 50μl of virus (2 x 10E3 TCID50/ml) in a 96-well plate at 37°C for 60min in 8 replicates. Hundred μl of Vero E6 cell (3 x 10E5 cells/ml) were then added to the mixture and incubated at 37°C, 5% CO2 until microscopy examination on day 4 to assess CPE. For viral load (VL) quantification, RNA extraction of the 8 pooled replicates of each XAV-19 dilution was performed with NucliSENS EasyMag (BioMerieux) according to manufacturer’s protocol. The relative VL were assessed from cycle threshold values for ORF1ab gene obtained by the TaqPath™ COVID-19 RT-PCR (ThermoFisher, Waltham, USA) and by linear regression in log10 copies/ml with a standard curve realized from a SARS-CoV-2 positive nasopharyngeal sample quantified by Droplet-Digital PCR (Bio-Rad).

## RESULTS

### Phase II/III

#### Demographic and patient characteristics

EUROXAV was initiated in 2021 and expected to enrol 870 patients. Yet, due to the very low enrolment rate, the study was unlikely to be completed in a reasonable timeline and was suspended on October 31, 2022. An interim blind analysis was performed in November 2022, and the results were shared with the Independent DSMB who recommended to stop the study. At that time, 293 patients with confirmed COVID-19 were screened. Eligible patients were randomized to receive XAV-19 (N=139) or placebo (N=140) (Figure 1). Final analyses were performed on target population defined as ITT patients having received at least one dose of treatment and fulfilling the main inclusion criteria, *i.e.* aged 18 or over, weighing between 40 and 120kg at the time of signing the informed consent, requiring or not low-flow oxygen therapy (≤6L/min) with WHO score at 2, 3 or 4, (129 and 130 patients in the XAV-19 and the placebo arms respectively). The demographic and clinical characteristic at baseline were similar between groups. The median age was 56.4 years and 54.5% were male patients. Percentage of patients with a WHO score of 2, 3 and 4 was 25.6, 45.7 and 28.7 in the XAV-19 arm and 25.4, 46.2, 28.5 in the placebo arm (Table 1). While comorbidities were slightly higher in the XAV-19 arm (62% in XAV-19 group vs 49 in placebo group), both groups were comparable for age, gender, BMI, and WHO score.

**Figure 1.**
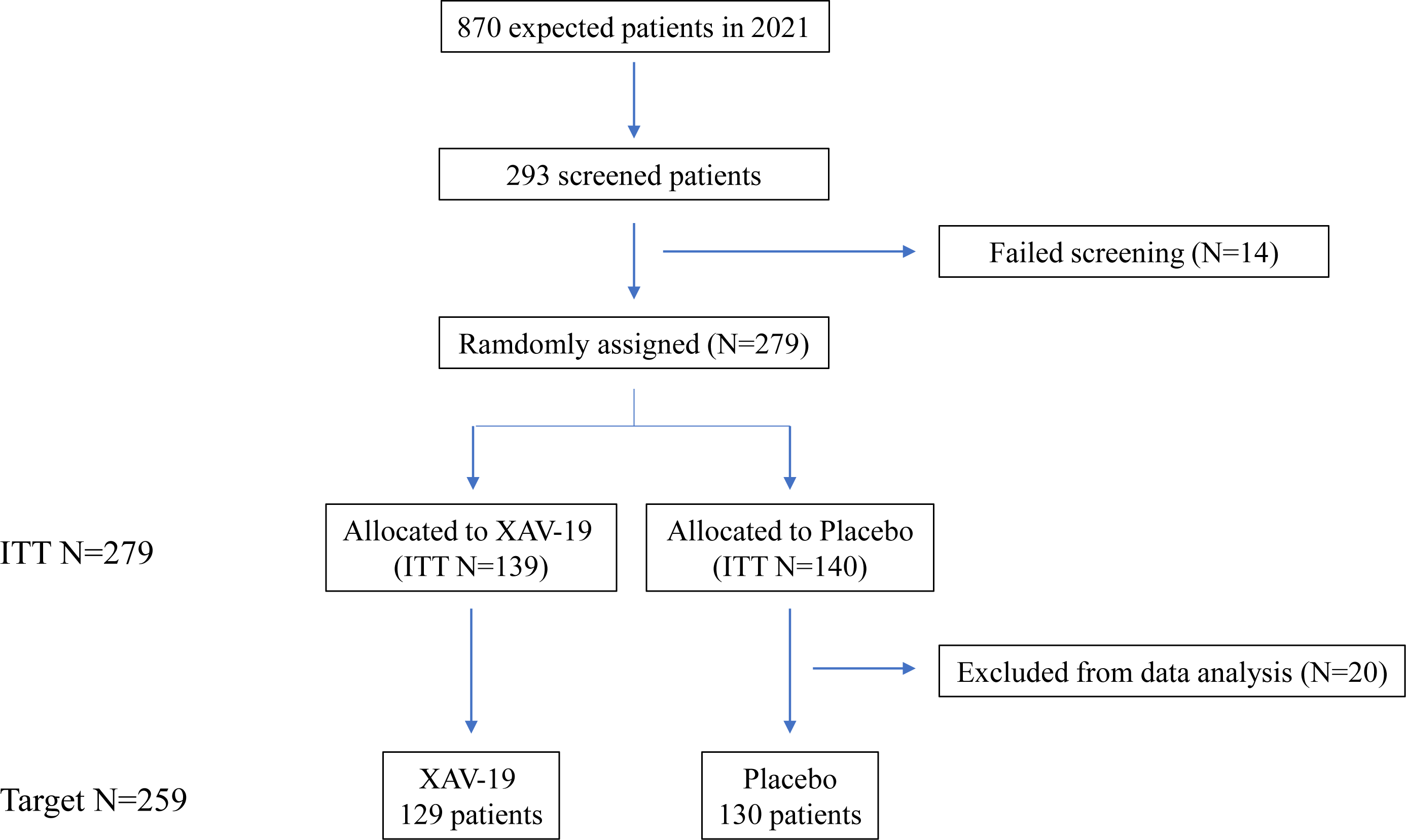
Clinical Trial Design.

**Table 1.** Demographic and baseline characteristics by treatment groups.

| Characteristic | XAV-19<br>(N=129) | PLACEBO<br>(N=130) | Total<br>(N=259) |
| --- | --- | --- | --- |
| Age at screening, mean (SD) | 57.5 (15.0) | 55.2 (15.4) | 56.4 (15.2) |
| Nb of male patients, n (%) | 74 (57.4) | 67 (51.5) | 141 (54.4) |
| BMI, mean (SD) | 28.3 (4.7) | 28.2 (4.6) | 28.2 (4.7) |
| Current smoker, n (%) | 14 (11.0) | 16 (12.5) | 30 (11.8) |
| WHO score at day 1, n (%) |  |  |  |
| WHO 2 | 33 (25.6) | 33 (25.4) | 66 (25.5) |
| WHO 3 | 59 (45.7) | 60 (46.2) | 119 (45.9) |
| WHO 4 | 37 (28.7) | 37 (28.5) | 74 (28.6) |
| Comorbidities<sup>§</sup>, n (%) | 80 (62.0) | 64 (49.2) | 144 (55.6) |
| Immunocompromised patients, n (%) | 11 (8.5) | 11 (8.5) | 22 (8.5) |
| Time between first COVID-19 symptoms and treatment in days, mean (SD) | 2.3 (2.3) | 2.6 (2.5) | 2.5 (2.4) |
| **Vital signs** |  |  |  |
| Systolic blood pressure, mmHg, mean (SD) | 125.1 (14.6) | 124.8 (16.3) | 124.9 (15.5) |
| Diastolic blood pressure, mmHg, mean (SD) | 76.5 (9.4) | 75.9 (9.8) | 76.2 (9.6) |
| Body temperature, mean (SD) | 37.3 (0.7) | 37.2 (0.8) | 37.3 (0.7) |
| **Respiratory examination** |  |  |  |
| Signs of pneumonia<sup>¥</sup>, n (%) | 122 (94.6) | 120 (93.0) | 242 (93.8) |
| Onset of the oldest COVID-19 symptom in days, mean (SD) | 5.5 (2.4) | 5.6 (2.6) | 5.5 (2.5) |
| SpO2 at ambient air (%), mean (SD) | 94.9 (1.8) | 94.7 (2.1) | 94.8 (1.9) |
| Patients under O2 supplementation, n (%) | 37 (28.7) | 37 (28.5) | 74 (28.6) |

¥: determined by chest X ray, CT scan or auscultation
§: BMI>30, diabetes DNID, diabetes DID, cardiac disorder, vascular disorder, hypercholesterolemia, renal failure, lung disease COPD and/or asthma

#### Primary efficacy endpoint

The primary endpoint was the proportion of patients with an aggravation of COVID-19 within 8 days after treatment. No differences on aggravation according to the treatment group was observed (12 *vs* 9 patients in the XAV19 and the placebo arm respectively, OR: 1.379, 95% CI 0.560 to 3.394, p=0.4831) (Table 2). Bivariate analyses showed that age at screening or comorbidities were associated with COVID-19 progression, while gender, BMI, or WHO score were not (Table 2). Logistic regression confirmed the association between age (over 70) and aggravation of COVID-19 (p=0.001, OR = 6.675 [2.158; 20.645]) (not shown).

**Table 2.** Bivariate analysis between categorical variables and aggravation of COVID-19.

| Parameters | No Worsening<br>N=238 | Worsening<br>N= 21 | Total<br>N=259 | P-values | OR<br>[95/% IC] |
| --- | --- | --- | --- | --- | --- |
| **Treatment, n (%)** |  |  |  |  |  |
| XAV19 | 117 (90.7) | 12 (9.3) | 129 | 0.4831 | 1.379 [0.560; 3.394] |
| PLACEBO | 121 (93.1) | 9 (6.9) | 130 |  |  |
| **Age at screening, n (%)** |  |  |  |  |  |
| ≤ 50 years | 87 (97.8) | 2 (2.2) | 89 | 0.0124 | 5.473 [1.245; 24.061] |
| > 50 years | 151 (88.8) | 19 (11.2) | 170 |  |  |
| ≤ 60 years | 144 (96) | 6 (4) | 150 | 0.0045 | 3.829 [1.435; 10.221] |
| > 60 years | 94 (86.2) | 15 (13.8) | 109 |  |  |
| ≤ 70 years | 196 (95.6) | 9 (4.4) | 205 | 0.0001 | 6.222 [2.464; 15.712] |
| > 70 years | 42 (77.8) | 12 (22.2) | 54 |  |  |
| ≤ 80 years | 230 (92.7) | 18 (7.3) | 248 | 0.0498 | 4.792 [1.169; 19.645] |
| > 80 years | 8 (72.7) | 3 (27.3) | 11 |  |  |
| **Gender, n (%)** |  |  |  |  |  |
| Male | 130 (92.2) | 11 (7.8) | 141 | 0.8433 | 0.914 [0.374; 2.233] |
| Female | 108 (91.5) | 10 (8.5) | 118 |  |  |
| **BMI, mean (SD)** | 28.2 (4.8) | 27.8 (2.6) | 28.2 |  |  |
| **Weight range n (%)** |  |  |  |  |  |
| Malnutrition : < 18.5 | 0 | 0 | 0 | 0.1561 | NA |
| Normal weight: [18.5;25[ | 65 (97) | 2 (3) | 67 |  |  |
| Overweight: [25;30[ | 92 (88.5) | 12 (11.5) | 104 |  |  |
| Moderate obesity: [30;35[ | 61 (92.4) | 5 (7.6) | 66 |  |  |
| Severe obesity: [35;40[ | 15 (6.3) | 0 (0) | 15 |  |  |
| Morbid or massive obesity: ≥ 40 | 5 (2.1) | 1 (5.0) | 6 |  |  |
| **WHO score at day 1, n (%)** |  |  |  |  |  |
| WHO 2/3 | 170 (91.9) | 15 (8.1) | 185 | 1 | 1.000 [0.372; 2.685] |
| WHO 4 | 68 (91.9) | 6 (8.1) | 74 |  |  |
| **Comorbidities, n (%)** |  |  |  |  |  |
| NO | 111 (96.5) | 4 (3.5) | 115 | 0.0147 | 3.715 [1.214; 11.368] |
| YES | 127 (88.2) | 17 (11.8) | 144 |  |  |

#### Secondary endpoints

There was no difference in the proportion of patients with COVID-19 aggravation within 8 days of treatment in the XAV-19 arm *vs* the placebo arm. Proportion of patients with COVID aggravation at day 15 was similar between groups. Proportion of patients with COVID-19 aggravation/improvement at day 15 *vs* day 8 was also similar between XAV-19 and placebo arms (not shown). Although proportions of aggravation/improvement were not different between groups, improvement did occur earlier in the XAV-19 arm (p=0.0340) (Figure 2A). This benefit was even more significant for non-hospitalized patients (WHO score of 2 at day 1) or patients not requiring supplemental oxygen (WHO score of 2 or 3 at day 1) (p=0.0003 and p=0.0159) while not significant for hospitalized patients (WHO score of 3 or 4) or hospitalized patients requiring supplemental oxygen (WHO score of 4) (p=0.7511 and p=0.8059). Furthermore, median time to improvement was reduced in the XAV-19 arm compared to the placebo one (4 days *vs* 14 days and 7 days *vs* 14 days for patients with a WHO score at baseline of 2 and 2/3 respectively) (Figure 2B). By contrast, time to aggravation was not different between XAV-19 and placebo groups (not shown).

**Figure 2.**
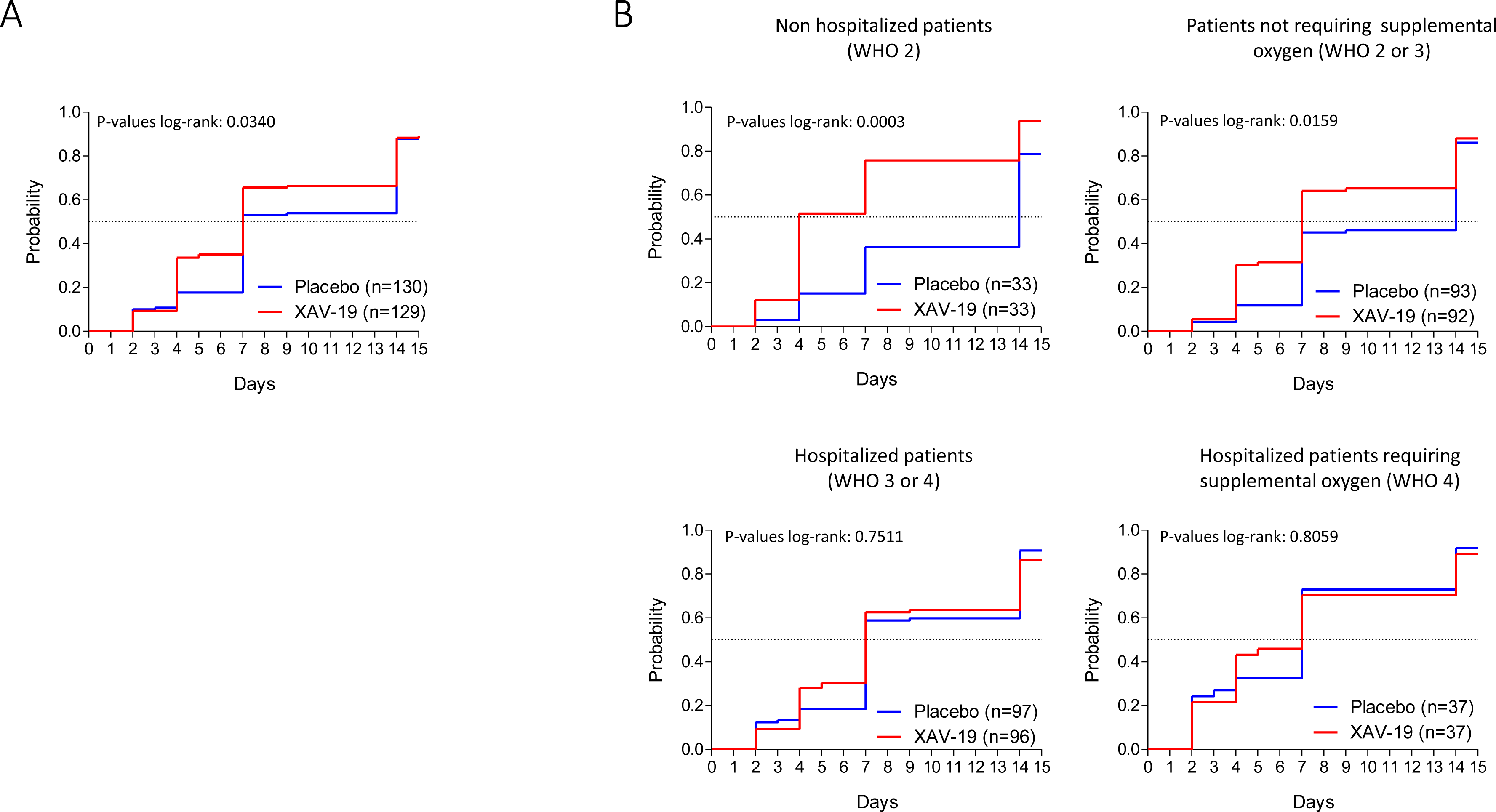
Time to clinical improvement. Kaplan-Meier curves by randomization group on whole TARGET population (**A**) or on TARGET population with a score of 2, 2 to 3, 3 to 4 or 4 at baseline (**B**). Improvement was defined as a diminution of at least 1 point on the WHO score compared to the WHO score at day 1. Dotted lines represent the median time to improvement.

#### Safety

A total of 83 adverse event (AE) (45 in the XAV-19 arm and 38 in the placebo arm) were observed from 29 patients in each group (Table 3). The percentage of mild/moderate AE in the XAV-19 arm was 91% (*vs* 76.3% in the placebo arm). Twenty-two severe AE (SAE) were reported in the XAV-19 group compared to 13 in the placebo group. None of them were considered as related by the investigators. Interestingly, SAE >3 account for 86% of SAE in the XAV-19 arm *vs* 92% in the placebo arms. Number of fatal SAE was equivalent between the two groups with 5 deaths in each group, from respiratory failure, multi-organ failure or disease progression. No anaphylaxis or hypersensitivity reactions and no infusion-related events were reported in any patients receiving XAV-19.

**Table 3.**
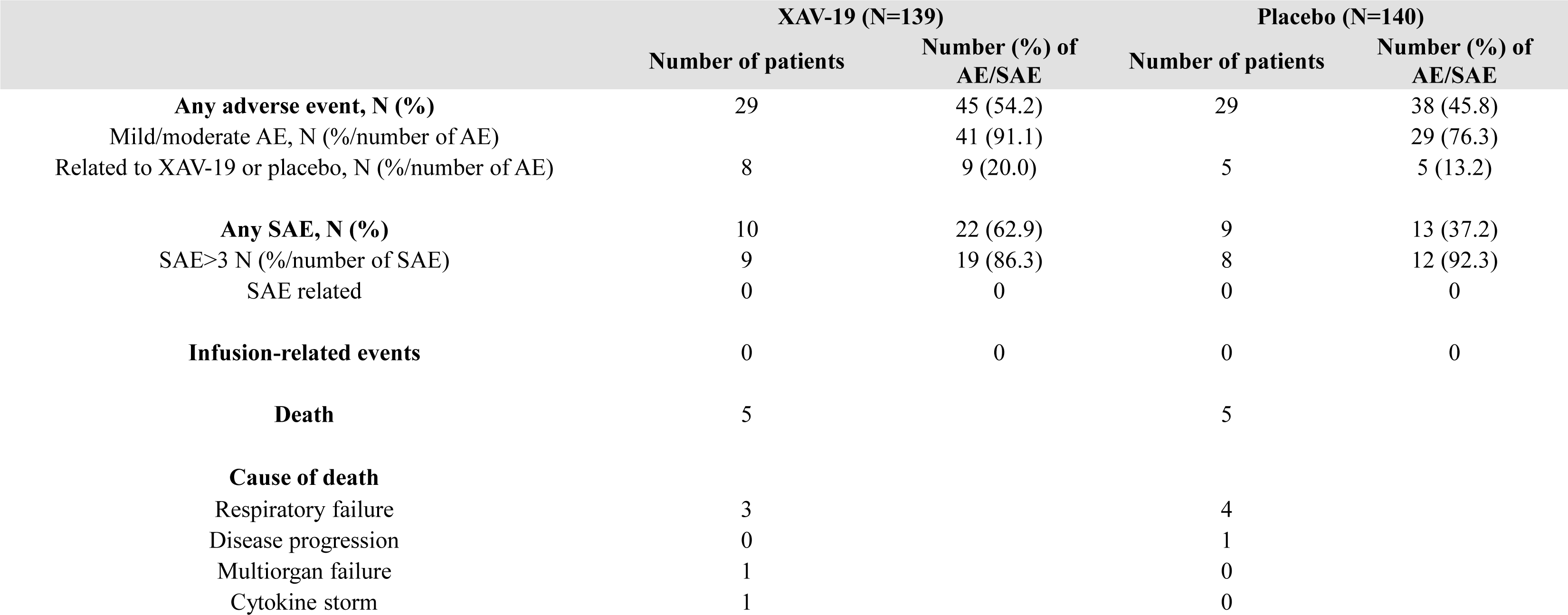
Adverse and serious adverse events.

To further explore XAV-19 clinical potential, the neutralizing activity of XAV-19 against Omicron and its subvariants was determined.

### In vitro antiviral activity

#### XAV-19 antiviral efficacy against Omicron and its subvariants

Binding of XAV-19 to Omicron, BA.2, BA2.12.1, BA.4/5 and BQ1.1 RBD appeared slightly lower than binding to the original strain RBD. Yet, similar plateau was reached whatever the variant tested (Figure 3A, top). In comparison, binding of the association tixagevimab/cilgavimab (Evusheld®) to Omicron or BA.4/5 RBD was dramatically reduced, and binding to BQ1.1 RBD was completely abolished. Although binding of BA2 and BA2.12.1 subvariants was less affected, a lower plateau was obtained (Figure 3A, bottom).

**Figure 3:**
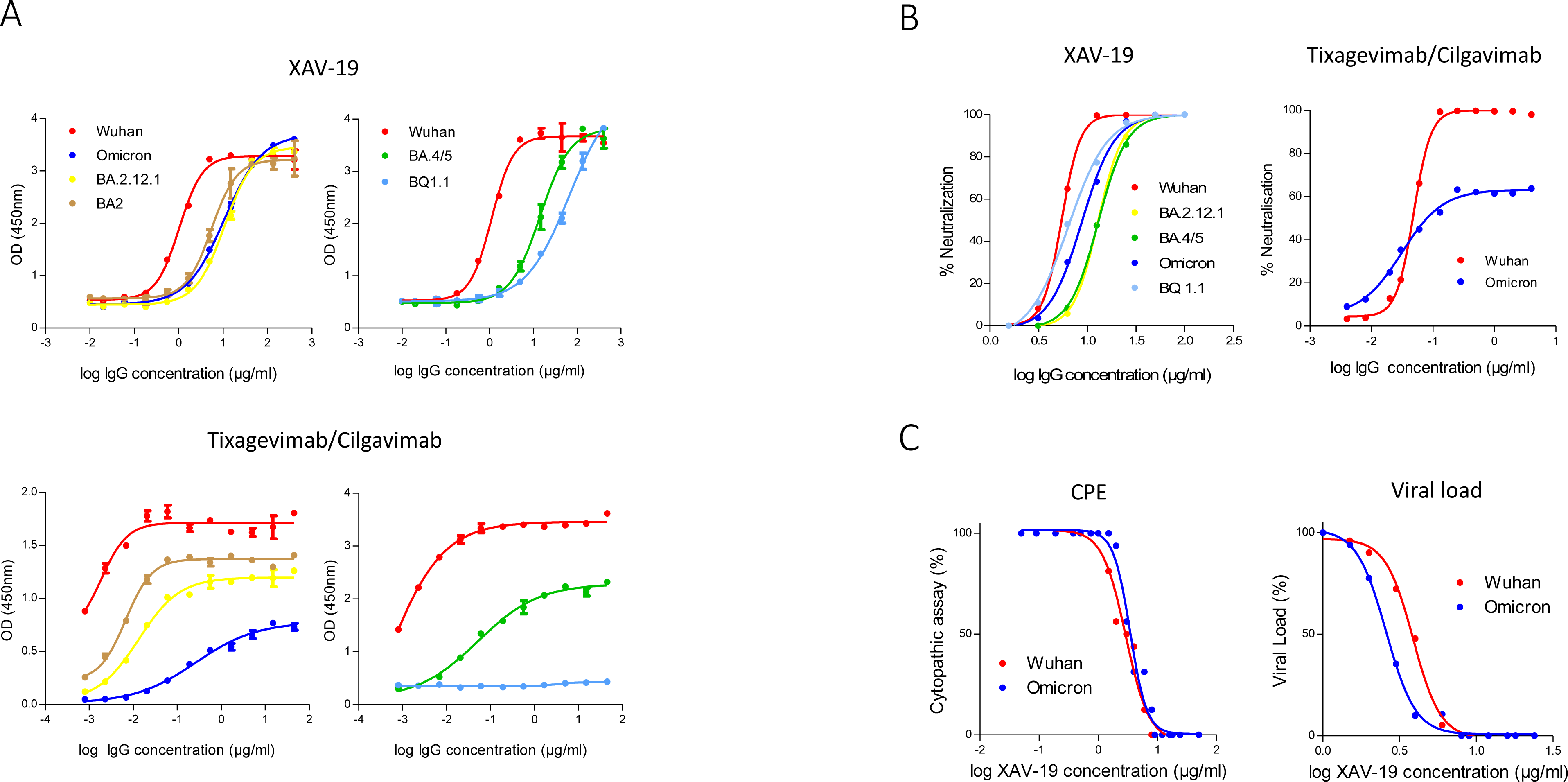
XAV-19 antiviral activity against Omicron and its subvariants. Binding of XAV-19 or Evusheld to Wuhan or Omicron and its subvariant RBD (**A**). XAV-19 or Evusheld were added to RBD coated plates at the indicated concentration and revealed with an HRP-conjugated secondary anti-pig or anti-human antibody. Neutralizing activity of XAV-19 or Evusheld to Wuhan or Omicron and its subvariant RBD (**B**). Recombinant His-Tag-RBD pre-incubated with XAV-19 or Evusheld were added to human ACE2 coated plates. Bound RBD were then detected with HRP-conjugated anti-His-Tag antibody. Neutralizing activity of XAV-19 on whole replicating viruses (**C**). Vero E6 cells were infected with Wuhan or Omicron SARS-CoV-2 strains. CPE was assessed by microscopy examination and viral load percentage was determined by quantitative RT-PCR.

XAV-19 neutralization potential was then tested in an RBD/ACE-2 binding competition assay. Comparable dose response profiles with full neutralization by XAV-19 were obtained with all the variants tested (Figure 3B, left); IC50 was moderately increased (5.9 ± 0.2 and 9.1 ±-0.6µg/mL for Wuhan and Omicron RBD respectively). By contrast, neutralization efficacy of tixagevimab/cilgavimab was limited to 60% for Omicron versus 100% for Wuhan RBD (Figure 3B, right). XAV-19 neutralization of viral infectivity was confirmed by CPE assay using SARS-CoV-2 clinical isolates. XAV-19 neutralized 100% of CPE for all the variants tested and reached 100% viral load reduction (Figure 3C) with slightly varying NT50 concentrations. Interestingly, NT50 against Omicron was in the low range in CPE and lower than for all other variants in VL assessment.

#### XAV-19 targets epitopes outside the Omicron mutations

Four main target epitopes of XAV-19 (347-fasvyawnr-417, 409-qiapgqtgn-417, 445-vsgnynylyrlfrksnlkpferdisteiy-473 and 530-stnlvk-535) were identified on Omicron RBD by proteolytic epitope mapping (Figure 4, bold), including 6 amino acids out of the 17 (in blue) directly involved in ACE-2 receptor binding. Among the 15 Omicron mutations in RBD (in yellow), only 2 lie inside the major XAV-19 target epitopes (N417 and S446) compared to 5 for the association tixagevimab/cilgavimab (underlined; K440, S446, N477, K478, A484)(16).

**Figure 4:**
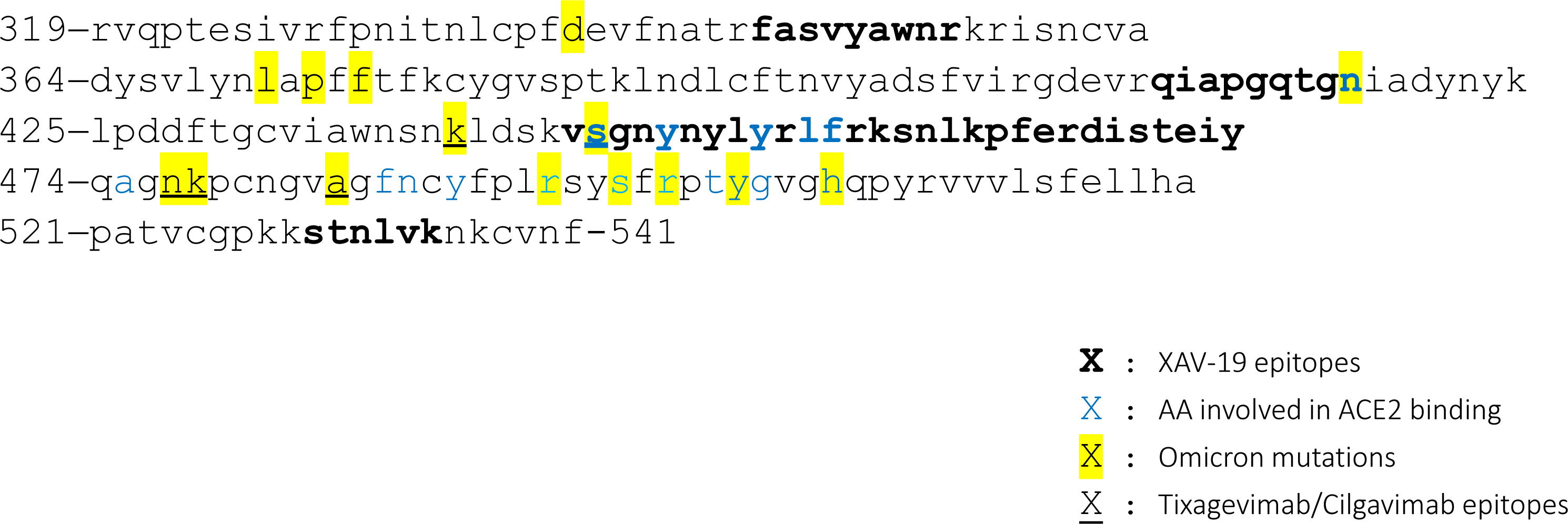
XAV-19 target epitopes lie outside the Omicron mutation sites. Amino acid sequence of the SARS-CoV-2 spike RBD variant Omicron and XAV-19 target epitopes (amino acid sequence numbered according to DBSOURCE sequence reference NC_045512.2). Bold: XAV-19 target epitopes confirmed by proteolytic epitope mapping; blue: amino acids in contact with ACE-2; yellow: mutations found in Omicron, differentiating from the original Wuhan RBD, underlined: Tixagevimab/Cilgavimab target epitopes.

## DISCUSSION

Although vaccination is now the primary option against SARS-CoV-2, therapeutic drugs are still needed for immunocompromised or at-risk individuals. The potential of anti-SARS-COV-2 mAb is today largely dampened by the continual emergence of resistant variants (4,5,17) and as SARS-CoV-2 will probably continue to mutate, robust and variant resistant treatments are still necessary.

The therapeutic potential of XAV-19 was investigated in the phase II/III EUROXAV study. Treatment with XAV-19 was safe and well tolerated. The fact that no patient who received XAV-19 had anaphylaxis or hypersensitivity reactions, confirms the validity of our glyco-humanization strategy. Yet, our trial suffered from a low inclusion rate and was stopped after inclusion of 293 patients. In addition, the proportion of worsening cases within the placebo arm was far lower than initially expected (8% observed *vs* 15% expected). This reduction in disease severity, although most patients were not vaccinated, might be due to the improvement in the management of COVID-19 patient (18,19) and/or to the infection of patients with the Omicron strain, known to be less pathogenic than the original one (20). Therefore, the low frequency of events, and the reduced recruitment rate, lowered dramatically the power of the study and the primary endpoint was not met. Nevertheless, we showed that XAV-19 efficiently modulated the kinetic of COVID-19 disease as it accelerated the clinical improvement of patients not requiring high-flow oxygen therapy (WHO score ≤ 4). More precisely, XAV-19 benefits to patients not requiring oxygen, while it does not bring any benefit to oxygen-dependent patients (WHO score 4). In that respect, our data match recent studies showing the suitability of therapeutics antibodies in patients with mild to moderate COVID-19 (21–23).

We have previously shown the neutralization efficiency of XAV-19, against the original Wuhan and the Alpha, Beta, Gamma and Delta variants (8). Here the neutralization activity of XAV-19 against Omicron and its subvariants was confirmed, while the association tixagevimab/cilgavimab (Evusheld®) lost its activity, as already published (17). Interestingly, XAV-19 even neutralized BQ1.1 subvariant, against which most mAb or cocktails of mAb are devoid of activity (17,24). Neutralization potency of XAV-19 was also confirmed on live Omicron by CPE assay. As the correlation between neutralization ELISA and CPE assay had been previously shown, (8), our data strongly suggest the ability of XAV-19 to neutralize live Omicron subvariants.

Due to its high number of mutations (30 mutations in the spike protein, 15 in the RBD), Omicron showed resistance to more than 80% of the therapeutic antibody candidate (25). Yet these mutations represent only 3% of the main target epitopes of XAV-19. The transmissibility of SARS-CoV-2 results from a fine balance between affinity to ACE-2 and capacity to escape immune response. The need to maintain sufficient affinity to ACE-2 probably limits SARS-CoV-2 RBD mutability as shown by the R493Q reversion mutation observed on the Omicron variants (26). Whereas antibody avidity allows to overcome SARS-CoV-2 variability (25), our data confirm that targeting multiple alternative epitopes, grants pAb to keep their binding and neutralization capacity (8). Thereby, the neutralizing activity of XAV-19, was confirmed so far on the 5 main SARS-CoV-2 variants that have emerged since the beginning of the pandemic.

Limitations of the study are the following. The 7-point ordinal scale WHO score as main endpoint, may suffer from discrepancies among practices in the different countries / hospitals; and standardized criteria for use of oxygen or hospitalization have not been implemented in this trial. Another limitation is the absence of information about vaccination though most patients were not vaccinated due to the main enrolment period from Jan to Sept 2021 (268 patients included) in locations where the vaccines were not available yet.

Altogether our data support XAV19 as a potent and affordable therapeutic option in patients with mild to moderate COVID-19, that could complement mAb antibody strategies. Today, absence of robust, variant resistant anti-SARS-COV-2 mAb therapy for immunocompromised individuals remains a public health concern (27). XAV-19 could be specially proposed for those high-risk patients.

## Data Availability

All data produced in the present study are available upon reasonable request to the authors

## Acknowledgments

We wish to thank all the patients, family members and staff from all the units that participated in the study.

## EUROXAV study group

Todor Atanasov, Dan Corneci, Valentin Cuervas-Mons, Gheorghe Lulian Diaconescu, Nikolay Evgeniev, Gerd Fätkenheuer, Elizabeth George, Rosen Georgiev, Benoit Guery, Guillen Santiago Moreno, Bedreag Ovidiu-Horea, Diamantis Kofteridis, Ali Mert, Symeon Metallidis, Hristo Metev, Cristina Mussini, Garyfallia Poulakou, Emmanuel Roilides, Fehmi Tabak, Pilar Vizcarra.

## References

1. DeWolf S, Laracy JC, Perales M-A, Kamboj M, van den Brink MRM, Vardhana S. SARS-CoV-2 in immunocompromised individuals. Immunity (2022) 55:1779–1798. doi: 10.1016/j.immuni.2022.09.006

2. Turtle L, Thorpe M, Drake TM, Swets M, Palmieri C, Russell CD, Ho A, Aston S, Wootton DG, Richter A, et al. Outcome of COVID-19 in hospitalised immunocompromised patients: An analysis of the WHO ISARIC CCP-UK prospective cohort study. PLoS Med (2023) 20:e1004086. doi: 10.1371/journal.pmed.1004086

3. Gupta A, Konnova A, Smet M, Berkell M, Savoldi A, Morra M, Van averbeke V, De Winter FHR, Peserico D, Danese E, et al. Host immunological responses facilitate development of SARS-CoV-2 mutations in patients receiving monoclonal antibody treatments. Journal of Clinical Investigation (2023) 133:1–13. doi: 10.1172/JCI166032

4. Focosi D, McConnell S, Casadevall A, Cappello E, Valdiserra G, Tuccori M. Monoclonal antibody therapies against SARS-CoV-2. Lancet Infect Dis (2022) 22:e311–e326. doi: 10.1016/S1473-3099(22)00311-5

5. Focosi D, Maggi F, Franchini M, McConnell S, Casadevall A. Analysis of immune escape variants from antibody-based therapeutics against covid-19: A systematic review. Int J Mol Sci (2022) 23: doi: 10.3390/ijms23010029

6. Widyasari K, Kim J. A Review of the Currently Available Antibody Therapy for the Treatment of Coronavirus Disease 2019 (COVID-19). Antibodies (2023) 12:5. doi: 10.3390/antib12010005

7. Cunha LER, Stolet AA, Strauch MA, Pereira VAR, Dumard CH, Gomes AMO, Monteiro FL, Higa LM, Souza PNC, Fonseca JG, et al. Polyclonal F(ab’)2 fragments of equine antibodies raised against the spike protein neutralize SARS-CoV-2 variants with high potency. iScience (2021) 24: doi: 10.1016/j.isci.2021.103315

8. Vanhove B, Marot S, So RT, Gaborit B, Evanno G, Malet I, Lafrogne G, Mevel E, Ciron C, Royer P-J, et al. XAV-19, a Swine Glyco-Humanized Polyclonal Antibody Against SARS-CoV-2 Spike Receptor-Binding Domain, Targets Multiple Epitopes and Broadly Neutralizes Variants. Front Immunol (2021) 12:1–11. doi: 10.3389/fimmu.2021.761250

9. Gaborit B, Dailly E, Vanhove B, Josien R, Lacombe K, Dubee V, Ferre V, Brouard S, Ader F, Vibet M, et al. Pharmacokinetics and Safety of XAV-19, a Swine Glyco-humanized Polyclonal Anti-SARS-CoV-2 Antibody, for COVID-19-Related Moderate Pneumonia: a Randomized, Double-Blind, Placebo-Controlled, Phase IIa Study. Antimicrob Agents Chemother (2021) 65:1–13. doi: 10.1128/AAC.01237-21

10. Lopardo G, Belloso WH, Nannini E, Colonna M, Sanguineti S, Zylberman V, Muñoz L, Dobarro M, Lebersztein G, Farina J, et al. RBD-specific polyclonal F(ab’)2 fragments of equine antibodies in patients with moderate to severe COVID-19 disease: A randomized, multicenter, double-blind, placebo-controlled, adaptive phase 2/3 clinical trial. EClinicalMedicine (2021) 34:100843. doi: 10.1016/j.eclinm.2021.100843

11. Taiwo BO, Chew KW, Moser C, Wohl DA, Daar ES, Li JZ, Greninger AL, Bausch C, Luke T, Hoover K, et al. Phase 2 safety and antiviral activity of SAB-185, a novel polyclonal antibody therapy for non-hospitalized adults with COVID-19. J Infect Dis (2023)1–6. doi: 10.1093/infdis/jiad013

12. Vanhove B, Duvaux O, Rousse J, Royer PJ, Evanno G, Ciron C, Lheriteau E, Vacher L, Gervois N, Oger R, et al. High neutralizing potency of swine glyco-humanized polyclonal antibodies against SARS-CoV-2. Eur J Immunol (2021) 51:1412–1422. doi: 10.1002/eji.202049072

13. Dhar C, Sasmal A, Varki A. From “Serum Sickness” to “Xenosialitis”: Past, Present, and Future Significance of the Non-human Sialic Acid Neu5Gc. Front Immunol (2019) 10:807. doi: 10.3389/fimmu.2019.00807

14. Vanhove B, Marot S, So RT, Gaborit B, Evanno G, Malet I, Lafrogne G, Mevel E, Ciron C, Royer P-J, et al. XAV-19, a Swine Glyco-Humanized Polyclonal Antibody Against SARS-CoV-2 Spike Receptor-Binding Domain, Targets Multiple Epitopes and Broadly Neutralizes Variants. Front Immunol (2021) 12:1–11. doi: 10.3389/fimmu.2021.761250

15. Cao B, Wang Y, Wen D, Liu W, Wang J, Fan G, Ruan L, Song B, Cai Y, Wei M, et al. A Trial of Lopinavir–Ritonavir in Adults Hospitalized with Severe Covid-19. New England Journal of Medicine (2020) 382:1787–1799. doi: 10.1056/nejmoa2001282

16. Dong J, Zost SJ, Greaney AJ, Starr TN, Dingens AS, Chen EC, Chen RE, Case JB, Sutton RE, Gilchuk P, et al. Genetic and structural basis for SARS-CoV-2 variant neutralization by a two-antibody cocktail. Nat Microbiol (2021) 6:1233–1244. doi: 10.1038/s41564-021-00972-2

17. Arora P, Kempf A, Nehlmeier I, Schulz SR, Jäck H-M, Pöhlmann S, Hoffmann M. Omicron sublineage BQ.1.1 resistance to monoclonal antibodies. Lancet Infect Dis (2023) 23:22–23. doi: 10.1016/S1473-3099(22)00733-2

18. Kumari M, Lu RM, Li MC, Huang JL, Hsu FF, Ko SH, Ke FY, Su SC, Liang KH, Yuan JPY, et al. A critical overview of current progress for COVID-19: development of vaccines, antiviral drugs, and therapeutic antibodies. J Biomed Sci (2022) 29:1–36. doi: 10.1186/s12929-022-00852-9

19. Brady DK, Gurijala AR, Huang L, Hussain AA, Lingan AL, Pembridge OG, Ratangee BA, Sealy TT, Vallone KT, Clements TP. A guide to <scp>COVID</scp> −19 antiviral therapeutics: a summary and perspective of the antiviral weapons against <scp>SARS-CoV</scp> −2 infection. FEBS J (2022)1–31. doi: 10.1111/febs.16662

20. Nyberg T, Ferguson NM, Nash SG, Webster HH, Flaxman S, Andrews N, Hinsley W, Bernal JL, Kall M, Bhatt S, et al. Comparative analysis of the risks of hospitalisation and death associated with SARS-CoV-2 omicron (B.1.1.529) and delta (B.1.617.2) variants in England: a cohort study. The Lancet (2022) 399:1303–1312. doi: 10.1016/S0140-6736(22)00462-7

21. Gupta A, Gonzalez-Rojas Y, Juarez E, Crespo Casal M, Moya J, Rodrigues Falci D, Sarkis E, Solis J, Zheng H, Scott N, et al. Effect of Sotrovimab on Hospitalization or Death Among High-risk Patients With Mild to Moderate COVID-19. JAMA (2022) 327:1236. doi: 10.1001/jama.2022.2832

22. Huang DT, McCreary EK, Bariola JR, Minnier TE, Wadas RJ, Shovel JA, Albin D, Marroquin OC, Kip KE, Collins K, et al. Effectiveness of Casirivimab-Imdevimab and Sotrovimab During a SARS-CoV-2 Delta Variant Surge. JAMA Netw Open (2022) 5:e2220957. doi: 10.1001/jamanetworkopen.2022.20957

23. O’Brien MP, Forleo-Neto E, Sarkar N, Isa F, Hou P, Chan KC, Musser BJ, Bar KJ, Barnabas R V., Barouch DH, et al. Effect of Subcutaneous Casirivimab and Imdevimab Antibody Combination vs Placebo on Development of Symptomatic COVID-19 in Early Asymptomatic SARS-CoV-2 Infection: A Randomized Clinical Trial. JAMA (2022) 327:432–441. doi: 10.1001/jama.2021.24939

24. Planas D, Bruel T, Staropoli I, Guivel-Benhassine F, Porrot F, Maes P, Grzelak L, Prot M, Mougari S, Planchais C, et al. Resistance of Omicron subvariants BA.2.75.2, BA.4.6, and BQ.1.1 to neutralizing antibodies. Nat Commun (2023) 14:824. doi: 10.1038/s41467-023-36561-6

25. Callaway HM, Hastie KM, Schendel SL, Li H, Yu X, Shek J, Buck T, Hui S, Bedinger D, Troup C, et al. Bivalent intra-spike binding provides durability against emergent Omicron lineages: Results from a global consortium. Cell Rep (2023) 42:112014. doi: 10.1016/j.celrep.2023.112014

26. Huo J, Dijokaite-Guraliuc A, Liu C, Zhou D, Ginn HM, Das R, Supasa P, Selvaraj M, Nutalai R, Tuekprakhon A, et al. A delicate balance between antibody evasion and ACE2 affinity for Omicron BA.2.75. Cell Rep (2023) 42:111903. doi: 10.1016/j.celrep.2022.111903

27. Casadevall A, Focosi D. SARS-CoV-2 variants resistant to monoclonal antibodies in immunocompromised patients constitute a public health concern. Journal of Clinical Investigation (2023) 133:1–3. doi: 10.1172/JCI168603

